# Preliminary Rasch analysis of the Multidimensional Assessment of Interoceptive Awareness in adults with stroke

**DOI:** 10.1101/2022.03.09.22272162

**Authors:** Jena Blackwood, Sydney Carpentier, Wei Deng, Ann Van de Winckel

## Abstract

**Purpose:** The Multidimensional Assessment of Interoceptive Awareness (MAIA) measures interoceptive body awareness, which includes aspects such as attention regulation, self-regulation, and body listening. Our purpose was to validate the MAIA in adults with stroke using Rasch Measurement Theory.

**Methods:** The original MAIA has 32 items grouped into 8 separately scored subscales that measure aspects of body awareness. Using Rasch Measurement Theory we evaluated the unidimensionality of the entire scale and investigated person and item fit, person separation reliability, targeting, local item dependence, and principal components analysis of residuals.

**Results:** Forty-one adults with chronic stroke (average 3.8 years post-stroke, 13 women, average age 57±13 years) participated in the study. Overall fit (χ^2^=62.26, *p*=0.26) and item fit were obtained after deleting 3 items and rescoring 26 items. One participant did not fit the model (2.44%). There were no floor (0.00%) or ceiling effects (0.00%). Local item dependence was found in 42 pairs. The person separation reliability was 0.91, and the person mean location was 0.06±1.12 logits.

**Conclusions:** The MAIA demonstrated good targeting and reliability, as well as good item and person fit in adults with chronic stroke. A study with a larger sample size is needed to validate our findings.

## Introduction

Interoception is a multifaceted construct that has inspired investigation and debate among numerous clinical professions since the early 20th century [1–3]. Charles Sherrington (1906) was the first to describe internal visceral physical processes as interoceptive, and external processing including processing pain, temperature, light, sound, and smell, as exteroceptive [3–6]. Since then, interoception has been recognized as a complex interplay of major biological systems that can broadly be defined as the conscious or unconscious process of perceiving, interpreting, integrating, and managing bodily signals such as pain and autonomic sensory information [1–3,6–9].

Therefore, the proposed taxonomy of interoceptive terms has expanded to encompass the multiple facets of interoception such as interoceptive accuracy (e.g., heartbeat detection accuracy), and self-reported interoceptive body awareness or interoceptive sensibility, which is self-reported sensory awareness in response to “physiological states, processes (including pain and emotion), and actions (including movement)” [1,5,6,10,11].

Historically, increased body awareness and focus on internal sensations were associated with hypochondria, catastrophizing, and maladaptive coping strategies [6,7]. Contemporary advocates of mind-body training and therapies promote mindful body awareness –a non-judgmental conscious moment-to-moment presence coupled with acceptance of information from the body– as beneficial to overall physical, mental, and emotional health and function [6,7,12–14].

The growing popularity of body awareness training practices such as yoga, Tai Chi, and Qigong in Western culture, coupled with a therapeutic interest in body awareness training and mindfulness practices, such as meditation and mindfulness-based stress reduction, has spurred investigations into the benefits of body awareness training and mindfulness practices across diverse clinical populations [7,10,12,15,16].

In persons with stroke, higher levels of body awareness have been associated with increased independence in performing activities of daily living, improved balance, and decreased fall risk [17]. In adults with stroke, yoga may be beneficial in regaining function and improving mental health, upper extremity strength, ambulation, balance, and achieving a greater connection and body acceptance [18–21]. Tai Chi has been shown to improve dual-task performance and reduce fall risk in individuals with stroke, while Qigong has shown reductions in cognitive decline [22–26].

Worldwide, approximately 15 million people suffer a stroke each year. Roughly, half are left with permanent disabilities impeding upper limb use in daily activities, balance, and ambulation as well as influencing mental health and coping strategies [27–35]. As part of the ongoing body of research, it is essential to use outcome measures that support accurate assessment of body awareness, especially when body awareness is assessed in studies with interventions aimed at training body awareness in this population.

Self-detection of interoceptive body awareness is commonly measured by self-report questionnaires, and a variety of them have been used to investigate changes in participants’ responses to body awareness training interventions [6,7,36]. In 2009, Mehling *et al*. explored the construct of body awareness and self-report measures available at that time and reported that the available measures were unable to differentiate between “non-judgmental, meditative, mindful awareness of body sensations” and “anxiety-related hyper-vigilance toward body sensations” [7].

At that time, Mehling *et al*. (2012) adopted an operational definition of body awareness as an iterative process encompassing sensory awareness stemming from physiological states including pain, emotion, and movement and includes cognitive appraisal shaped by attitudes, culture, and individual belief systems [6]. Moreover, “dimensions of critical importance” were created to assist in recognizing the multifaceted nature of the construct’s modern understanding [6].

Subsequently, in 2012, Mehling *et al*. developed the MAIA as a subjective measure of body awareness through an extensive mixed-methods process including focus group development of new scale items to fit the defined construct. Items were then field-tested with expert and novice mind-body practitioners [6,7,15].

The MAIA is a 32-item self-report instrument consisting of eight subscales spanning 5 overall dimensions and 13 subdimensions of interoceptive body awareness. More details on these dimensions and subdimensions can be found in Mehling *et al*. (2012) [6]. Items are scored on a Likert scale between 0 = Never to 5 = Always with unlabeled interim integers. Each of the 8 subscales is scored separately by averaging the items belonging to the respective subscale. A higher score on each subscale indicates “more positive body awareness”, and though the Not-Distracting and Not-Worrying scales contain reverse-scored items, the higher scores reflect less worrying and less distraction when faced with discomfort [6,37]. In its original version, a total MAIA score is not calculated [6,38,39].

The MAIA was intended to differentiate between experienced and inexperienced mind-body practitioners, to measure changes in participants’ outcomes after mind-body therapies, and to “differentiate between anxiety-driven hypervigilance and mindful attention styles” [6,11,40]. As such, the MAIA has been heavily utilized for body awareness training measurement in varied populations, but to the best of the authors’ knowledge, it has been used in only one previous observational study with adults with stroke [35,39]. At the time of publication, it has been translated into over 24 languages [41].

Internal consistency, correlations with related construct measures and between subscales, and confirmatory factor analysis were performed on the MAIA during its initial publication [6]. Mehling *et al*. (2012) originally demonstrated that most MAIA subscales had moderate to strong correlations with each other, but the “Not Distracting” and “Not Worrying” subscales showed only small to moderate correlations to the other subscales [6,39]. The internal consistency, as measured by Cronbach’s alpha, was below 0.70 for the subscales “Noticing”, “Not-Distracting”, and “Not-Worrying” and was confirmed in some subsequent studies. The other subscales ranged from 0.79 to 0.87 [6,37,42–54]. The confirmatory factor analysis confirmed the multidimensionality of the scale [6].

Other researchers have since discarded the negatively worded items, changed the phrasing to positively worded sentences, or made other changes to the scale to improve the factor loading of the scale during some of the translations and validation studies [43–45,48,51,54–56]. Ferenzti *et al*. (2020) found that all the MAIA items measured “a general interoceptive awareness factor” with confirmatory factor analysis, but this general factor did not correlate with the subscales “Not-Distracting” and “Not-Worrying” [39].

Rasch Measurement Theory (RMT) can be used to assess the unidimensionality and the structural validity of a scale. RMT is a probabilistic model stating persons with a higher interoceptive body awareness would have a higher probability of obtaining a higher score on the items. RMT can therefore be used to verify whether the items fit the probabilistic mathematical Rasch model. With this analysis, the scale is transformed from an ordinal to an interval measurement which improves instrument precision [57–63]. Structural validity and unidimensionality are evaluated through the overall fit of the scale, the item and person fit, threshold order of scoring categories within each item, person separation reliability (PSR), targeting, floor and ceiling effects, mean error variance, principal components analysis of residuals (PCAR), and local item dependence (LID) with residual correlations [59,60].

The aim of this study was to perform a Rasch analysis of the MAIA to evaluate its structural validity in adults with stroke and to assess its unidimensionality. To the best of our knowledge, the MAIA has not been validated with RMT before. Therefore, we performed a Rasch analysis to assess the structural validity of the MAIA in individuals with stroke for use in future research and clinical applications with this population.

## Methods

### Participants

Potential participants contacted the Brain Body Mind Lab members after seeing the study flier on the University Campus and in clinics, or through postings on the university websites. We included adults between 18-99 years of age who had an ischemic or hemorrhagic stroke and were medically stable, English speaking, and able to consent. We excluded participants with cognitive impairments (Mini-mental State Exam-brief version, <13/16), contractures in the upper extremity that would hinder testing arm movements, severe aphasia or apraxia, or other medical conditions that would preclude participation in the study. The study was approved by the University of Minnesota’s Internal Review Board (IRB#STUDY00000821). The study was performed in accordance with the Declaration of Helsinki. All participants signed informed consent on paper and filled out the MAIA on paper.

### Data Collection

We collected information on age, the hemispheric side of the stroke, time after stroke, and whether the stroke was ischemic or hemorrhagic. We assessed pain with the Numeric Pain Rating Scale (NPRS), depression with the Patient Health Questionnaire (PHQ-9), and stroke-related neurologic deficit with the NIH stroke scale (NIHSS) [64–66]. Participants also indicated if they had taken part in any breathing exercises in the past if they were currently participating in breathing exercises regularly, and if they had past body awareness training experiences such as dance, martial arts, Tai Chi, Qigong, yoga, Pilates, or other body awareness training.

### Statistical Analysis

Rasch analysis was performed using the Rasch Unidimensional Measurement Model (RUMM) 2030 software (RUMM Laboratory, Perth, WA) using a partial credit model and rating scale polytomous model to analyze overall fit, scoring thresholds, individual and item fit, and overall fit with item-trait interactions. All reverse scoring as necessitated by the MAIA scoring protocol was completed before the Rasch analysis so that higher scores would imply higher interoceptive awareness across all items. We decided to investigate the unidimensionality of the MAIA by analyzing all items together rather than perform a Rasch analysis per identified subscale.

A “threshold” is the point at which adjacent categories have the same likelihood of being selected [67,68]. Disordered thresholds reveal that the logical estimated order of the scale construct is not measured appropriately by the item response categories and that respondents may have had difficulty differentiating between categories and/or that not all categories were used [60,68–72]. Fit statistics indicate the appropriateness of person and item fit to the Rasch model [68]. Person and item fit residuals greater than +2.50 indicate misfitting items or persons, and less than -2.50 indicate redundant items. Significant *p*-values for item fit are calculated with Bonferroni correction [60].

PSR differentiates the person’s ability levels of the trait for research or clinical purposes, with PSR > 0.90 allowing the researcher or clinician to make decisions for individuals and ≥ 0.70 to make group decisions [60,73,74]. Floor and ceiling effects above 15% are considered problematic [60, 72]. The average person location within a range of -0.50 and 0.50 logits of average item location indicates adequate assessment targeting [60,75]. PCAR assesses the random variance in residuals, and if the variance is due to the underlying trait rather than other components [60,76]. LID is found when an item pair shares a greater degree of content compared to other assessment items [77,78]. Standard residual correlations of 0.20 or greater than the average of the standard residual item correlation indicate the presence of LID [60,79].

## Results

Forty-one adults (average age 57±13.65 years, average time post-stroke 3.79±2.85 years) with unilateral ischemic brain lesions resulting in upper limb impairment participated in the study between September 27, 2017, and February 28, 2020. Participant demographics and stroke characteristics are detailed in Table 1.

**Table 1.** Demographic and clinical characteristics of study participants.

|  | Adults with stroke<br>(n=41) |
| --- | --- |
| Age (mean ± standard deviation; range) | 57±13 (25 – 81) |
| Sex |  |
| Men | 28 |
| Women | 13 |
| Type of Stroke |  |
| Ischemic | 31 |
| Hemorrhagic | 9 |
| Unknown | 1 |
| Hemisphere of Stroke |  |
| Left | 29 |
| Right | 12 |
| Time since stroke in years (mean; range) | 3.79 (.50 - 10.50) |
| Current participation in breathing exercises (yes; no) | 17; 24 |
| Past participation in breathing exercises (yes; no) | 18; 23 |
| Past participation in body awareness training (yes; no) | 19; 22 |
| Current pain (NPRS) (yes; no) | 21; 20 |
| Depression (PHQ-9 >10) (yes; no) | 6; 35 |
| NIHSS (mean; range) | 2.78; (0 - 9) |
| MMSE (mean; range) | 15; (13 - 16) |

Legend: NPRS = Numeric Pain Rating Scale, PHQ-9 = Patient Health Questionnaire–9, NIHSS = National Institutes of Health Stroke Scale Score, MMSE = Mini-Mental State Examination

The iteration table (S1 Table) details the step-by-step process of RMT analysis for the MAIA. More detailed explanations are below.

Upon initial analysis, 29 of the 32 items on the MAIA presented with disordered thresholds. Over the course of 5 iterations of rescoring, all 29 items were rescored by collapsing response categories. Nine of the original items were collapsed to dichotomous response categories [0 0 0 0 1 1], with the other rescored items varying on the spectrum of 3 to 4 collapsed response categories ([0 0 0 0 1 2], [0 0 0 1 1 2], [0 0 0 1 2 3], [0 0 1 1 2 2], [0 0 1 2 2 3]). Only items 14 and 15 retained the original scoring format [0 1 2 3 4 5].

Following rescoring, item 5 “*I do not notice (I ignore) physical tension or discomfort until they become more severe*.*”* displayed misfit (Fit Residual = 4.19; *p* = 0.0001) and was subsequently removed. Next, item 16 *“I can maintain awareness of my whole body even when a part of me is in pain or discomfort*.*”* and item 23 *“When I feel overwhelmed, I can find a calm place inside*.*”* were removed because they demonstrated a Guttman-like response pattern covering a wide logit-range from -9 logits to 1 logit as displayed (S1 Fig, S2 Fig). Next, items 11, 12, 18, 20, 26, 30, and 31 all displayed small thresholds in the middle scoring categories of each item’s scale and were rescored to improve fit to the model as indicated in the iteration table (S1 Table). Finally, item 25 was rescored due to a small threshold in the middle scoring categories. The resulting item location in logits for all items, after the above steps were completed, is listed in the S2 Table.

The person mean location was 0.05±1.13 logits indicating that the MAIA item difficulty was well-targeted for this population. There was no floor (0.00%) or ceiling effect (0.00%). We found that only 1 of the 41 participants displayed misfit (2.44% of the total group). The PSR was 0.91 indicating that the scale can be used for individual decision-making. The mean error variance was 0.12 logits, which is a small error estimate.

The revised Rasch-based MAIA is shown in Table 2 with items listed in order of difficulty on the logit scale from the easiest item at the top to the most difficult item at the bottom.

**Table 2.**
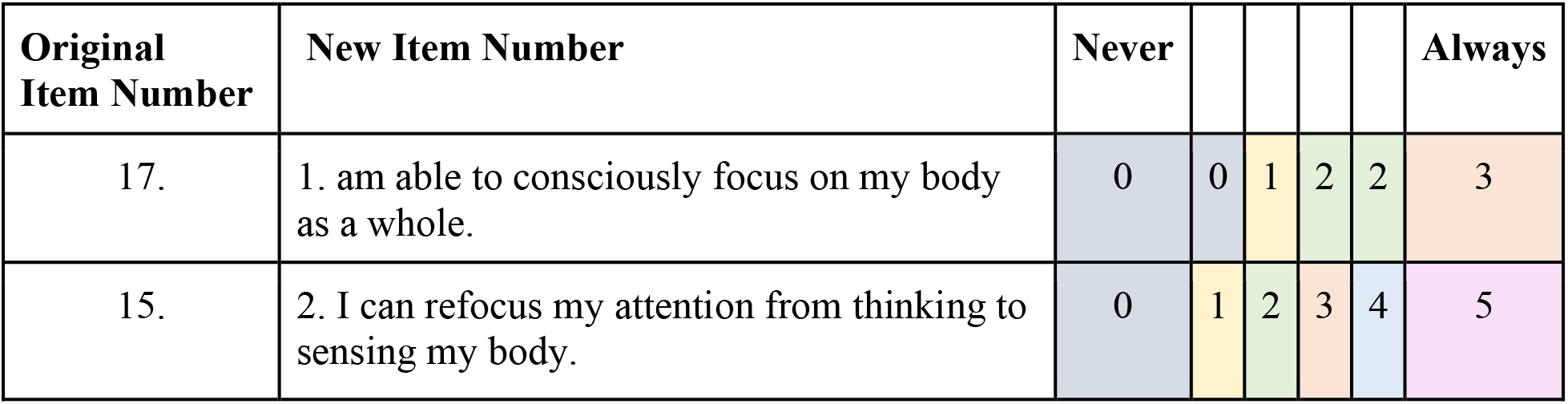
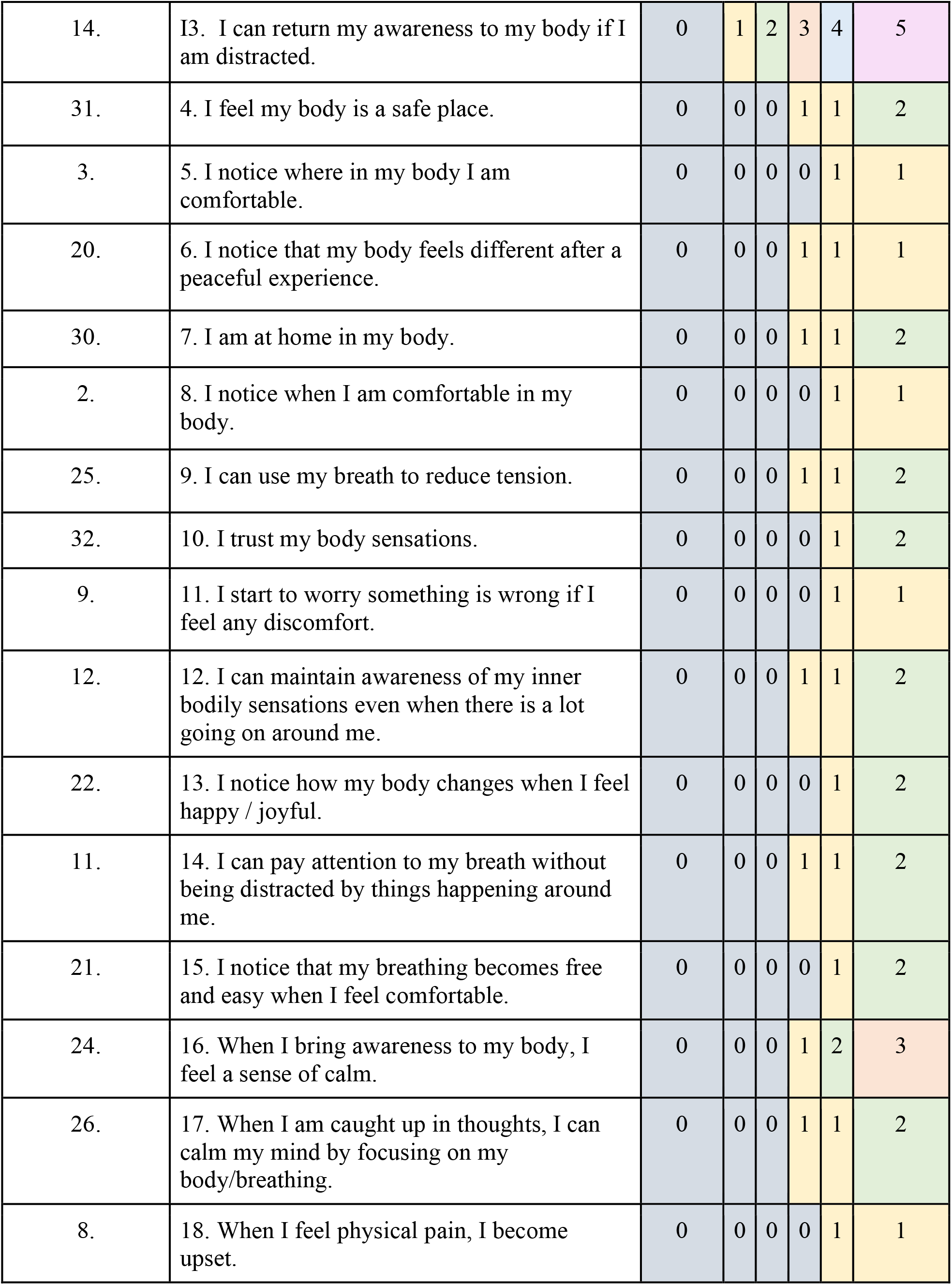
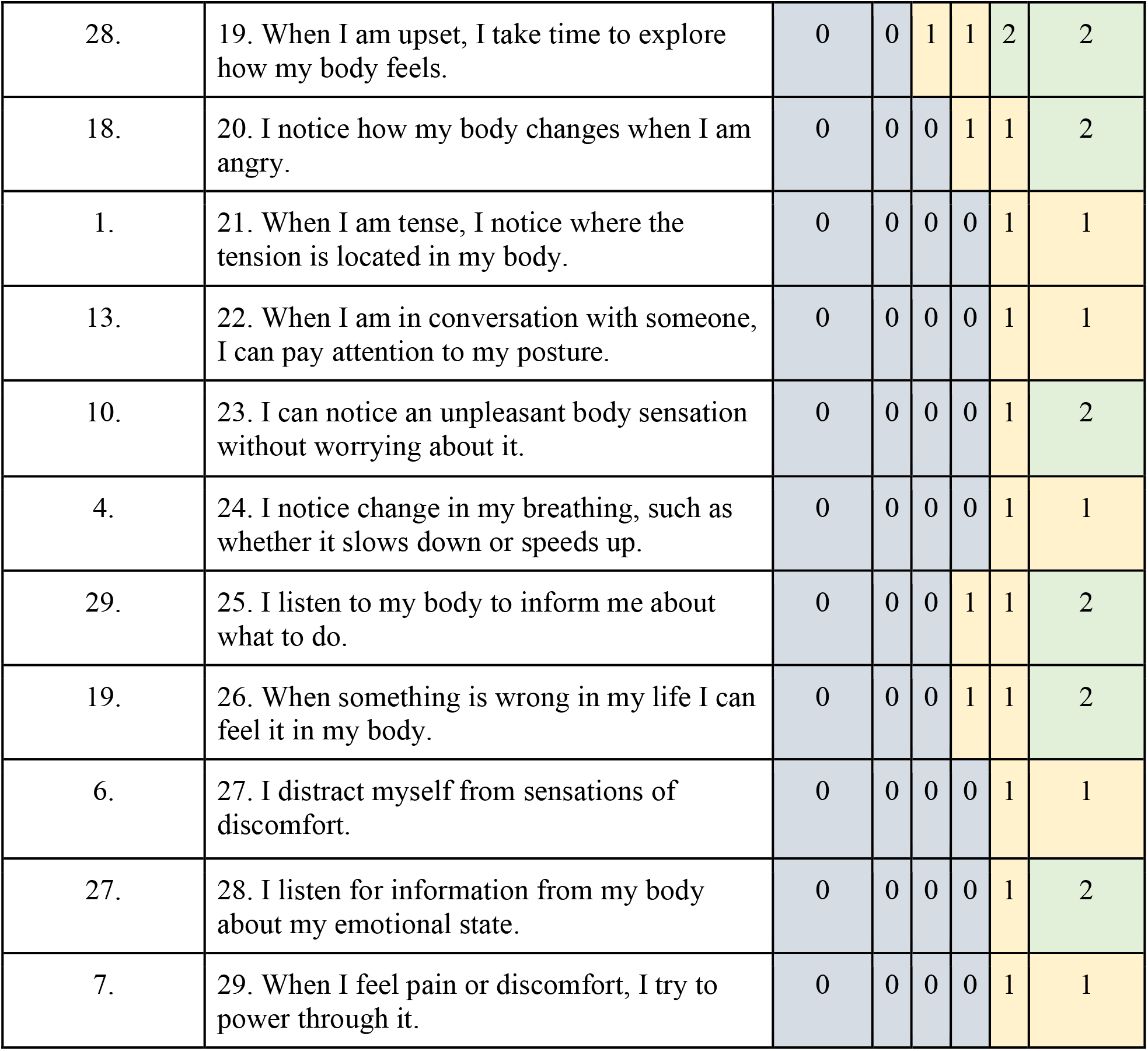
Rasch-based MAIA.

In sum, the Rasch analysis resulted in a 29-item MAIA scale with a good overall fit, and good item and person fit (S3 Fig).

We report the score-to-measure in Table 3, which shows the conversion from ordinal total scores to logits to logit-based % from 0-100 for the total score. Our score-to-measure data provided should only be used with full data sets.

**Table 3.** Rasch based MAIA score conversion.

| Total MAIA Score | Logit | Converted to logits<br>0-100 |
| --- | --- | --- |
| 0 | -5.93 | 0.00 |
| 1 | -5.00 | 8.66 |
| 2 | -4.33 | 14.95 |
| 3 | -3.84 | 19.51 |
| 4 | -3.45 | 23.12 |
| 5 | -3.13 | 26.09 |
| 6 | -2.86 | 28.60 |
| 7 | -2.63 | 30.76 |
| 8 | -2.42 | 32.68 |
| 9 | -2.24 | 34.39 |
| 10 | -2.07 | 35.94 |
| 11 | -1.92 | 37.37 |
| 12 | -1.77 | 38.69 |
| 13 | -1.64 | 39.91 |
| 14 | -1.52 | 41.07 |
| 15 | -1.40 | 42.16 |
| 16 | -1.29 | 43.20 |
| 17 | -1.18 | 44.20 |
| 18 | -1.08 | 45.16 |
| 19 | -0.98 | 46.08 |
| 20 | -0.88 | 46.97 |
| 21 | -0.79 | 47.83 |
| 22 | -0.70 | 48.69 |
| 23 | -0.61 | 49.52 |
| 24 | -0.52 | 50.33 |
| 25 | -0.43 | 51.13 |
| 26 | -0.35 | 51.93 |
| 27 | -0.26 | 52.72 |
| 28 | -0.18 | 53.51 |
| 29 | -0.09 | 54.30 |
| 30 | -0.01 | 55.09 |
| 31 | 0.08 | 55.89 |
| 32 | 0.16 | 56.68 |
| 33 | 0.25 | 57.49 |
| 34 | 0.34 | 58.31 |
| 35 | 0.43 | 59.14 |
| 36 | 0.52 | 59.99 |
| 37 | 0.61 | 60.86 |
| 38 | 0.71 | 61.74 |
| 39 | 0.81 | 62.65 |
| 40 | 0.91 | 63.59 |
| 41 | 1.01 | 64.56 |
| 42 | 1.12 | 65.57 |
| 43 | 1.23 | 66.62 |
| 44 | 1.35 | 67.73 |
| 45 | 1.48 | 68.89 |
| 46 | 1.61 | 70.14 |
| 47 | 1.75 | 71.46 |
| 48 | 1.91 | 72.90 |
| 49 | 2.08 | 74.47 |
| 50 | 2.26 | 76.22 |
| 51 | 2.48 | 78.20 |
| 52 | 2.73 | 80.51 |
| 53 | 3.03 | 83.32 |
| 54 | 3.42 | 86.96 |
| 55 | 3.99 | 92.26 |
| 56 | 4.82 | 100.00 |

The overall fit of the model χ^2^(DF)=74.59 (58), *p*=0.07, as well as item fit showed that the MAIA can assess interoceptive awareness as a unidimensional construct. The PCAR analysis showed an eigenvalue of 4.78 and a percentage variance of 16.49%, indicating that underneath the broader aspect of interoceptive awareness, the MAIA may encompass about 5 aspects of interoceptive awareness. The paired *t*-tests showed that the person location on the subtests created by the positive (items 2, 19, 20, 21, 22, 27, 28, 29) and negative (items 11, 12, 13, 14, 17, 30, 31) loadings on the first principal component showed that 21.95% of the person locations were significantly different between the subtests, confirming the underlying aspects of the MAIA. In the same vein, LID was found in 42 item pairs (S3 Table). The “Attention Regulation” and “Emotional Awareness” scales each contributed 14 pairs to LID.

## Discussion

The main results of this pilot Rasch analysis in adults with stroke showed a 29-item MAIA scale with a good overall fit, and good item and person fit (S3 Fig). Moreover, the MAIA was well-targeted for adults with chronic stroke. This result was obtained after extensive rescoring of the items, which may be explained for several different reasons.

First, when the MAIA items were developed, the original wording of question items asked respondents to rate “how true” an item was for them. However, after some pretests, this wording was found to be “difficult” and subsequently changed to “how often” this item was true for them [6]. Therefore, it makes sense that 9 of the 29 items we retained were maximally efficiently rescored in a dichotomous format, more like “True” or “False” questions. Additionally, using END form formatting (i.e., labeling extreme categories with unlabeled interim options) may have contributed to the necessity to merge several scoring categories [80,81].

There is also debate in the research community as to whether combining positively and negatively worded items in the same scale may result in measurement inaccuracies [82,83]. In our analysis, only 1 of the deleted reverse-scored items was negatively worded (item 5). However, several previous studies have omitted various negatively worded items from the “Not-Distracting” and “Not-Worrying” subscales for poor factor loading [42,43,45,47,51,54].

Our results confirmed that the MAIA measures approximately 5 overarching aspects of interoceptive awareness as originally found by Mehling *et al*. (2012). The items that had a positive loading on the first principal component came primarily from the “Emotional Awareness” (EA) (items 19, 20, 21, 22) and “Body Listening” (BL) (items 27, 28, 29) subscales with one item from the “Noticing” subscale (item 2). This fits with the original finding from Mehling *et al*. (2012) of moderate correlations (*r*=.60) between EA and BL scales. Mehling *et al*. (2012) also specified that items from the EA and BL subscales were part of the same overarching dimension of “Mind Body Integration” (dimension 5) whereas item 2 (“I notice where I am tense in my body.”) is from the dimension of “Noticing” (dimension 1) also originally known as “Awareness of Body Sensations” [6]

The items with negative loadings on the first principal component contained primarily items from the “Attention Regulation” (AR) (items 11, 12, 13, 14, 17) and “Trusting” (TR) (items 30, 31) subscales. Mehling *et al*. (2012) originally found the AR and TR scales to also be moderately correlated (*r*=.50) [6]. The AR and TR subscales were originally found to measure 2 different areas of interoceptive awareness, the “Capacity to Regulate Attention” (dimension 3) and “Trusting Body Sensations” (dimension 4) aspects respectively [6]. In other words, our analysis of positive and negative loadings is consistent with 4 of the original dimensions purported to be measured by the MAIA excluding only the dimension of “Emotional Reactions and Attentional Response to a Sensation ‘‘ which contains all the originally reverse-scored items from the “Not-Worrying” and Not-Distracting” subscales. This overarching dimension may be measured by the MAIA, yet we cannot confirm this based on our analysis.

To the best of our knowledge, only one other previous study utilized the MAIA in adults with stroke, specifically to measure recovery of body awareness impairments after acute stroke recovery, and to identify the associations between body awareness impairments and “sensation, motor impairment, self-efficacy, and quality of life” after stroke [35]. Serrada *et al*. (2021) found that body awareness as measured by the MAIA had a “poor association” with other measures utilized in their study including the NIHSS, the Functional Independence Measure, the Body Perception Disturbance scale, and the Stroke Impact Scale [64,84–87]. However, Serrada *et al*. (2021) did not have their study participants fill out the MAIA at baseline due to concerns of the MAIA being “inappropriate” to administer immediately after a stroke. Their participants were enrolled 1 - 14 days post-stroke, so consequently participants only filled out the MAIA at 1- and 6 months post-stroke. Serrada *et al*. (2021) did not recommend using the MAIA in adults after stroke due to the potential of the questions being “too distressing for someone in the acute stroke phase” and because their authors posited that neglect and hypo-vigilance would be more common after stroke than hypervigilance [35].

Serrada *et al*. (2021) proposed that the development of a measure of body awareness that is appropriate for use after acute stroke is necessary, and we agree. We did not encounter distress when acquiring the MAIA in our sample, but we recruited only adults with chronic stroke. Also, about 50% of our participants had past or current experience with breathing exercises and/or past body awareness training and thus the type of questions asked on the MAIA might have been more familiar to some of them.

### Study limitations

Our pilot Rasch results in our small sample are promising, but a RMT in a larger sample size is needed to confirm our findings. Also, we only recruited adults with chronic stroke and thus cannot generalize our findings to adults with acute or subacute stroke. A larger sample will also allow for DIF analysis as well as other types of reliability and reproducibility.

## Conclusion

Our pilot Rasch-based MAIA indicates promising results for future use of the MAIA in adults with chronic stroke. Further studies are needed to validate the findings and complete the psychometrics on the MAIA.

## Data Availability

The dataset supporting the conclusions of this article is available in the Data Repository for U of M (DRUM).

https://doi.org/10.13020/pgc5-6659

## Acknowledgments

The authors thank all participants as well as the volunteers who assisted the principal investigator (AVDW) at the Minnesota State Fair and Highland Fest. Our profound gratitude goes to Marc Noël for the critical review of the manuscript.

## Authors’ contributions

All authors contributed substantially to parts of the manuscript, critically revised it for content, approved the final version, and agreed to be accountable for the accuracy and integrity of this work. Specific contributions include:

- Conception or design of the work: AVDW
- Acquisition and analysis of evidence: AVDW, JB
- Interpretation of evidence: AVCW, JB, WD, SC
- Writing - original draft – AVDW, JB
- Writing - review and editing - AVDW, JB, WD, SC

## Supporting Information

**S1 Fig.**
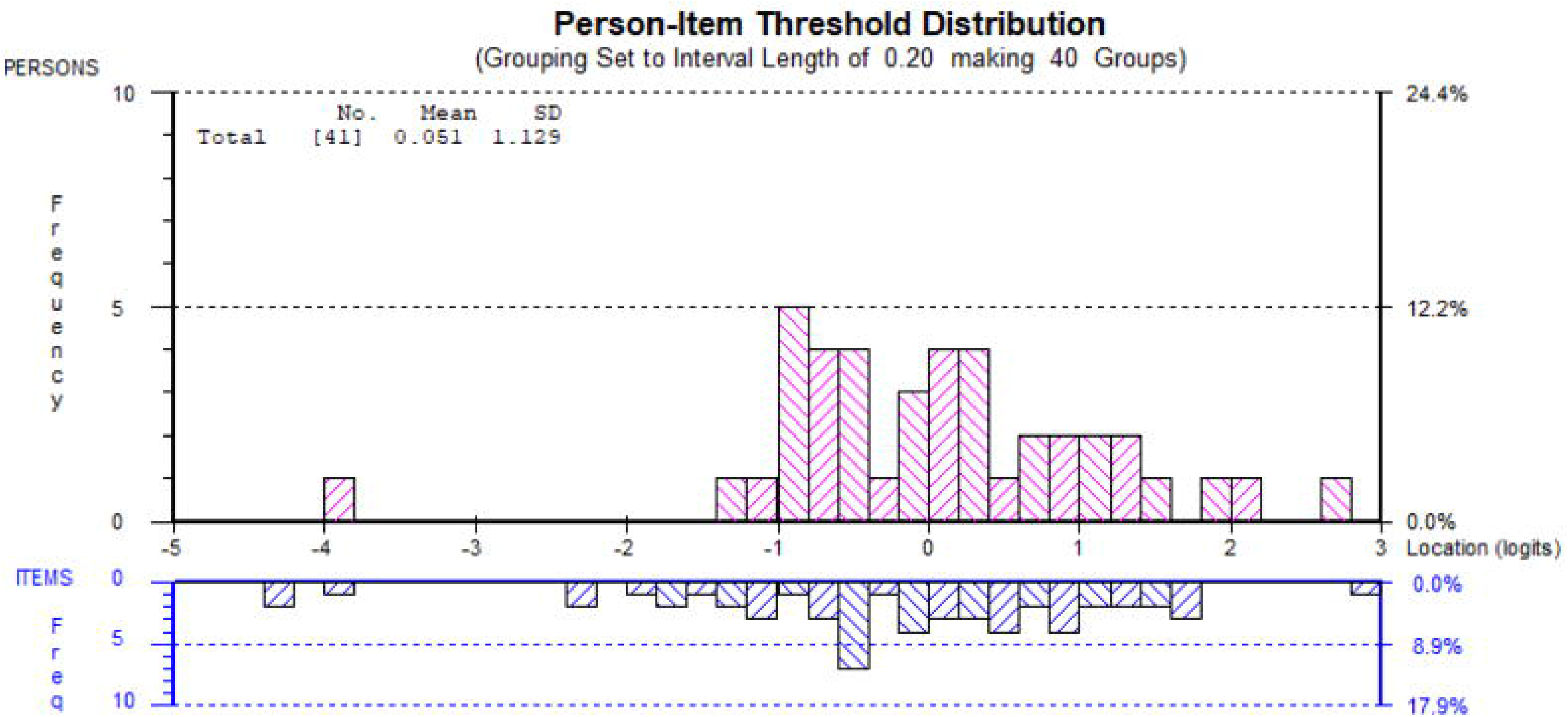
Category probability curve for Item 16 in adults with stroke. The category probability curve shows the probability of each category being selected on the Y-axis and the X-axis shows the item measured in logits demonstrating the person’s ability of their body awareness in relation to the question *“I can maintain awareness of my whole body even when a part of me is in pain or discomfort*.*”*

**S2 Fig.**
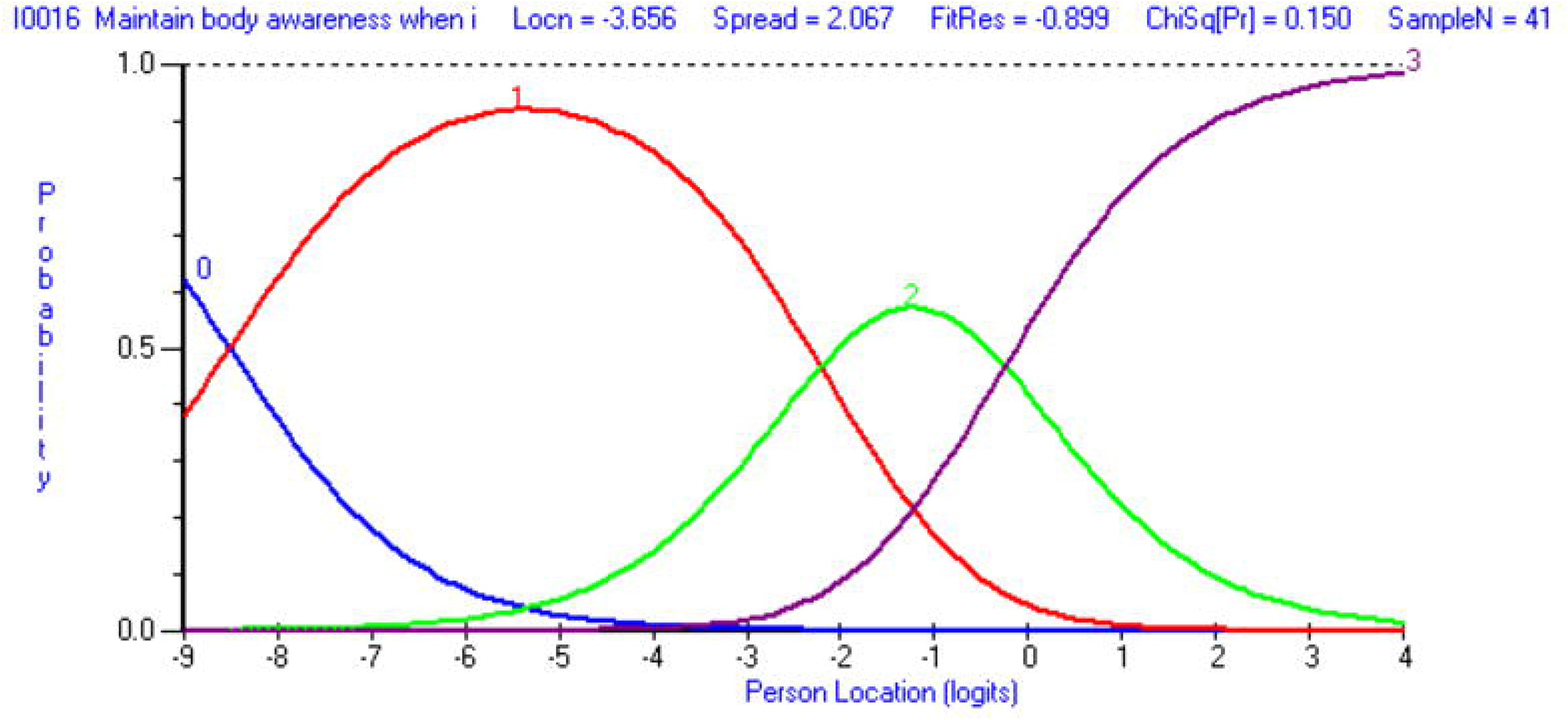
Category probability curve for Item 23 in adults with stroke. The category probability curve shows the probability of each category being selected on the Y-axis and the X-axis shows the item measured in logits demonstrating the person’s ability of their body awareness in relation to the question *“When I feel overwhelmed, I can find a calm place inside*.*”*

**S3 Fig.**
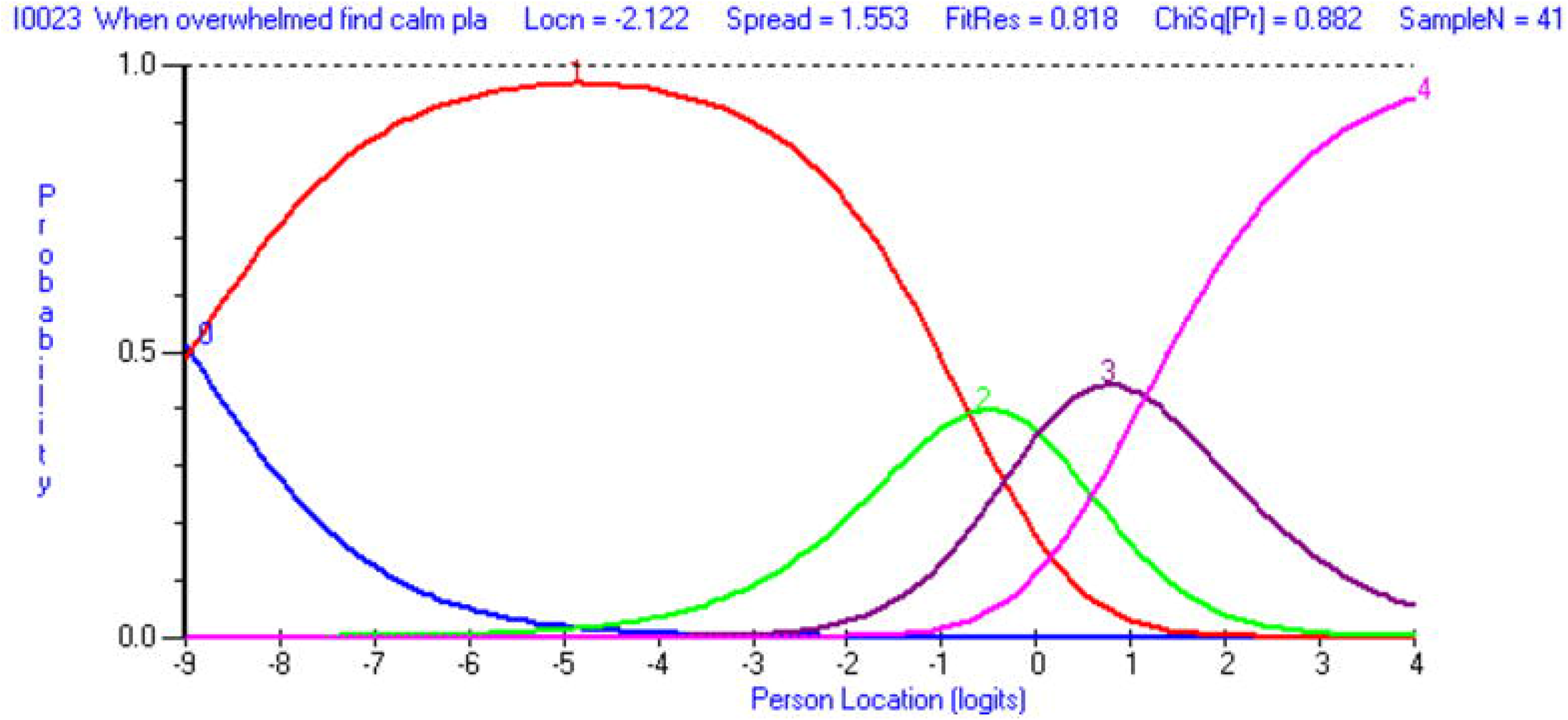
Person-item threshold distribution in adults with stroke. The X-axis logit ruler represents item difficulty and respondent ability. The blue histograms show item difficulty level frequencies, whereas the pink histograms represent the frequencies of the respondent’s ability level of interoceptive body awareness. Higher levels of body awareness are indicated by higher logit values.

**Table S1.**
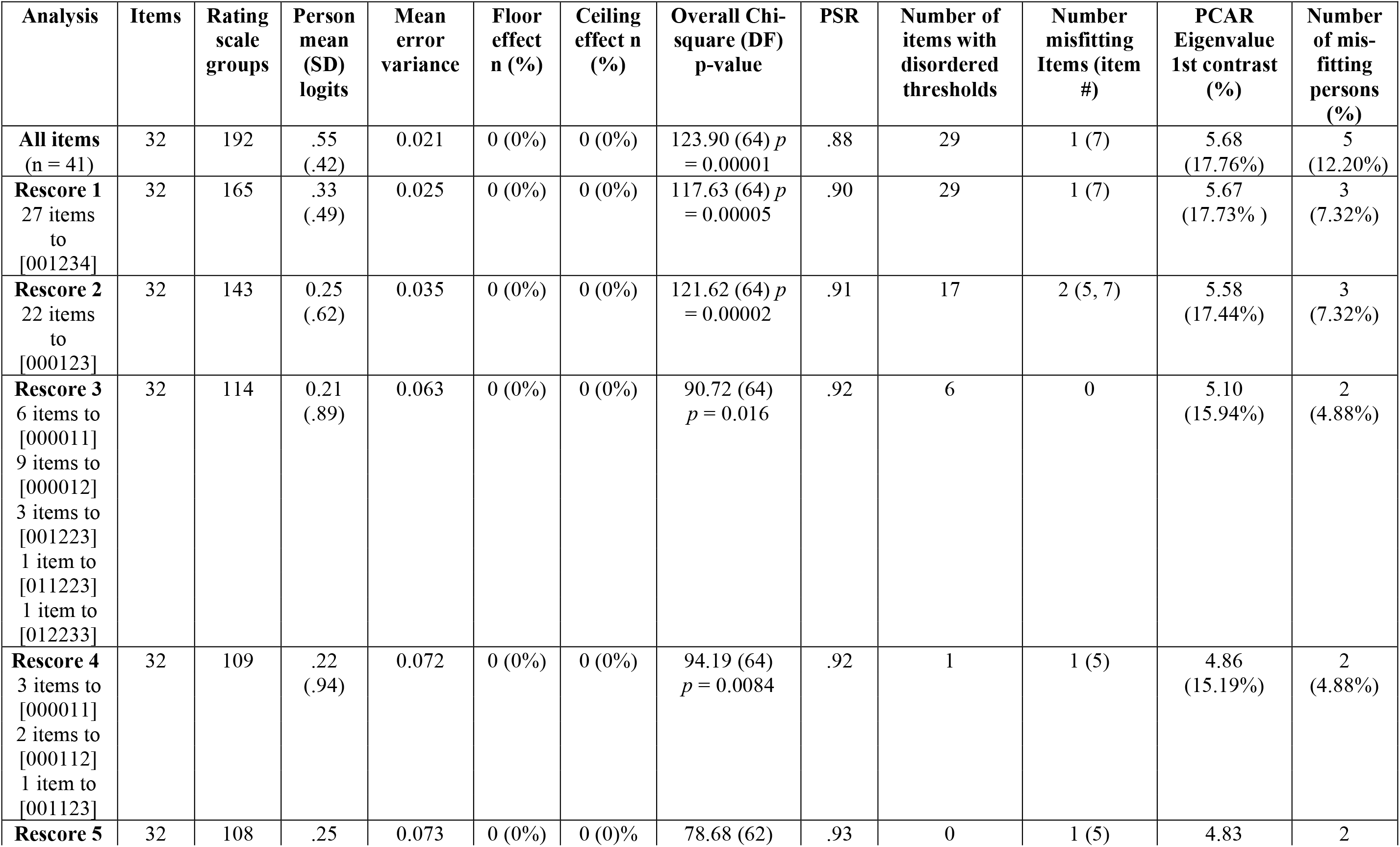
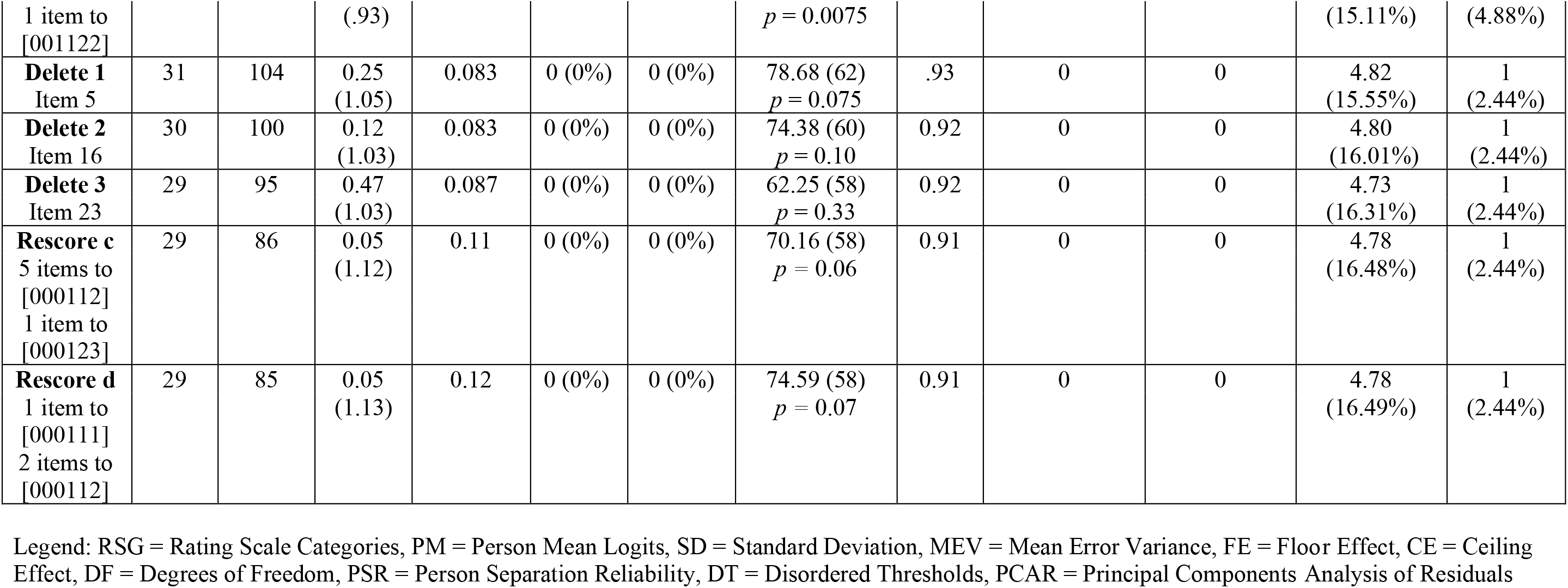
Iteration table MAIA.

**S2 Table.** Item fit statistics of the Rasch-based MAIA.

| **Original Item Number** | **Item number and Description of Rasch-based MAIA** | **Item location (logits)** | **SE** | **Fit Residuals** | **Chi-Square (DF=2)** | ***p*-value** |
| --- | --- | --- | --- | --- | --- | --- |
| Item 17 | 1. I am able to consciously focus on my body as a whole. | -1.63 | 0.26 | 0.27 | 3.13 | 0.21 |
| Item 15 | 2. I can refocus my attention from thinking to sensing my body. | -1.62 | 0.18 | 0.22 | 1.18 | 0.55 |
| Item 14 | 3. I can return awareness to my body if I am distracted. | -1.26 | 0.17 | 1.62 | 0.25 | 0.88 |
| Item 31 | 4. I feel my body is a safe place. | -0.83 | 0.29 | -0.54 | 1.06 | 0.59 |
| Item 3 | 5. I notice where in my body I am comfortable. | -0.48 | 0.35 | -0.11 | 0.02 | 0.99 |
| Item 20 | 6. I notice that my body feels different after a peaceful experience. | -0.48 | 0.35 | -0.19 | 0.67 | 0.72 |
| Item 30 | 7. I am at home in my body. | -0.47 | 0.28 | 0.00 | 0.81 | 0.67 |
| Item 2 | 8. I notice when I am comfortable in my body. | -0.37 | 0.35 | -0.75 | 0.85 | 0.65 |
| Item 25 | 9. I can use my breath to reduce tension. | -0.29 | 0.29 | -0.46 | 0.51 | 0.78 |
| Item 32 | 10. I trust my body sensations. | -0.29 | 0.23 | -1.03 | 3.43 | 0.18 |
| Item 9 | 11. I start to worry that something is wrong if I feel any discomfort. | -0.19 | 0.35 | 2.45 | 7.79 | 0.02 |
| Item 12 | 12. I can maintain awareness of my inner bodily sensations even when there is a lot going on around me. | -0.16 | 0.27 | -0.40 | 3.17 | 0.21 |
| Item 22 | 13. I notice how my body changes when I am angry. | -0.13 | 0.24 | -0.75 | 1.24 | 0.54 |
| Item 11 | 14. I can pay attention to my breath without being distracted by things happening around me. | -0.05 | 0.25 | 0.50 | 1.03 | 0.60 |
| Item 21 | 15. I notice that my breathing becomes free and easy when I am comfortable. | -0.02 | 0.24 | 0.07 | 0.63 | 0.73 |
| Item 24 | 16. When I bring awareness to my body, I feel a sense of calm. | -0.01 | 0.21 | -1.67 | 6.18 | 0.05 |
| Item 26 | 17. When I am caught up in thoughts, I can calm my mind by focusing on my body/breathing. | 0.08 | 0.27 | -0.75 | 2.07 | 0.36 |
| Item 8 | 18. When I start to feel physical pain, I become upset. | 0.11 | 0.35 | 1.33 | 0.83 | 0.66 |
| Item 28 | 19. When I am upset, I take time to explore how my body feels. | 0.18 | 0.25 | 0.32 | 4.19 | 0.12 |
| Item 18 | 20. I notice how my body changes when I am angry. | 0.25 | 0.28 | -0.35 | 0.56 | 0.76 |
| Item 1 | 21. When I am tense, I notice where the tension is located in my body. | 0.26 | 0.35 | 0.12 | 0.12 | 0.94 |
| Item 13 | 22. When I am in conversation with someone, I can pay attention to my posture. | 0.35 | 0.35 | 0.77 | 2.17 | 0.34 |
| Item 10 | 23. I can notice an unpleasant body sensation without worrying about it. | 0.37 | 0.24 | 3.10 | 3.47 | 0.18 |
| Item 4 | 24. I notice changes in my breathing, such as whether it slows down or speeds up. | 0.48 | 0.35 | 0.12 | 1.09 | 0.58 |
| Item 29 | 25. I listen to my body to tell me what to do. | 0.59 | 0.27 | 0.02 | 0.26 | 0.88 |
| Item 19 | 26. When something is wrong in my life, I can feel it in my body. | 0.62 | 0.27 | 0.21 | 3.08 | 0.21 |
| Item 6 | 27. I distract myself from sensations of discomfort. | 1.02 | 0.37 | 0.84 | 11.68 | 0.00 |
| Item 27 | 28. I listen for information from my body about my emotional state. | 1.09 | 0.26 | 0.31 | 11.55 | 0.46 |
| Item 7 | 29. When I feel pain or discomfort, I try to power through it. | 2.86 | 0.59 | 0.61 | 11.61 | 0.11 |

Legend: SE = Standard Error, DF = Degrees of Freedom

**S3 Table.** Local Item Dependence (Residual correlation ≥ 0.37)

| Item number | RC | Scale |
| --- | --- | --- |
| 1. When I am tense, I notice where the tension is located in my body. | 0.53 | NOT |
| 2. I notice when I am uncomfortable in my body. |  | NOT |
| 1. When I am tense, I notice where the tension is located in my body. | 0.42 | NOT |
| 3. I notice where in my body I am comfortable. |  | NOT |
| 2. I notice when I am uncomfortable in my body. | 0.47 | NOT |
| 3. I notice where in my body I am comfortable. |  | NOT |
| 1. When I am tense, I notice where the tension is located in my body. |  | NOT |
| 26. When I am caught up in my thoughts, I can calm my mind by focusing on my body/breathing. |  | SR |
| 8. When I feel physical pain, I become upset. | 0.49 | NW |
| 9. I start to worry something is wrong if I feel any discomfort. |  | NW |
| 11. I can pay attention to my breath without being distracted by things happening around me. | 0.59 | AR |
| 12. I can maintain awareness of my inner bodily sensations even when there is a lot going on around me. |  | AR |
| 12. I can maintain awareness of my inner bodily sensations even when there is a lot going on around me. | 0.58 | AR |
| 27. I listen for information from my body about my emotional state. |  | BL |
| 12. I can maintain awareness of my inner bodily sensations even when there is a lot going on around me. | 0.40 | AR |
| 29. I listen to my body to inform me about what to do. |  | BL |
| 13. When I am in conversation with someone, I can pay attention to my posture. | 0.55 | AR |
| 14. I can return awareness to my body if I am distracted. |  | AR |
| 13. When I am in conversation with someone, I can pay attention to my posture. | 0.39 | AR |
| 17. I am able to consciously focus on my body as a whole. |  | AR |
| 13. When I am in conversation with someone, I can pay attention to my posture. | 0.44 | AR |
| 19. When something is wrong in my life, I can feel it in my body. |  | EA |
| 13. When I am in conversation with someone, I can pay attention to my posture. | 0.38 | AR |
| 29. I listen to my body to inform me about what to do. |  | BL |
| 14. I can return awareness to my body if I am distracted. | 0.50 | AR |
| 17. I am able to consciously focus on my body as a whole. |  | AR |
| 14. I can return awareness to my body if I am distracted. | 0.41 | AR |
| 18. I notice how my body changes when I am angry. |  | EA |
| 14. I can return awareness to my body if I am distracted. | 0.42 | AR |
| 19. When something is wrong in my life, I can feel it in my body. |  | EA |
| 14. I can return awareness to my body if I am distracted. | 0.44 | AR |
| 29. I listen to my body to inform me about what to do. |  | BL |
| 15. I can refocus my attention from thinking to sensing my body. | 0.38 | AR |
| 24. When I bring awareness to my body, I feel calm. |  | SR |
| 17. I am able to consciously focus on my body as a whole. | 0.37 | AR |
| 19. When something is wrong in my life, I can feel it in my body |  | EA |
| 17. I am able to consciously focus on my body as a whole. | 0.48 | AR |
| 27. I listen for information from my body about my emotional state. |  | BL |
| 18. I notice how my body changes when I am angry. | 0.37 | EA |
| 19. When something is wrong in my life, I can feel it in my body. |  | EA |
| 18. I notice how my body changes when I am angry. | 0.47 | EA |
| 30. I am at home in my body |  | TR |
| 18. I notice how my body changes when I am angry. | 0.42 | EA |
| 31. I feel my body is a safe place |  | TR |
| 19. When something is wrong in my life, I can feel it in my body. | 0.51 | EA |
| 22. I notice how my body changes when I am angry. |  | EA |
| 20. I notice that my body feels different after a peaceful experience. | 0.51 | EA |
| 22. I notice how my body changes when I am happy / joyful. |  | EA |
| 20. I notice that my body feels different after a peaceful experience. | 0.39 | EA |
| 27. I listen for information from my body about my emotional state. |  | BL |
| 20. I notice that my body feels different after a peaceful experience. | 0.54 | EA |
| 28. When I am upset, I take time to explore how my body feels. |  | BL |
| 20. I notice that my body feels different after a peaceful experience. | 0.37 | EA |
| 30. I am at home in my body. |  | TR |
| 20. I notice that my body feels different after a peaceful experience. | 0.55 | EA |
| 31. I feel my body is a safe place. |  | TR |
| 21. I notice that my breathing becomes free and easy when I feel comfortable. | 0.39 | EA |
| 28. When I am upset, I take time to explore how my body feels. |  | BL |
| 21. I notice that my breathing becomes free and easy when I feel comfortable. | 0.43 | EA |
| 29. I listen to my body to inform me about what to do. |  | BL |
| 21. I notice that my breathing becomes free and easy when I feel comfortable. | 0.46 | EA |
| 30. I am at home in my body. |  | TR |
| 22. I notice how my body changes when I am angry. | 0.39 | EA |
| 30. I am at home in my body. |  | TR |
| 22. I notice how my body changes when I am angry. | 0.46 | EA |
| 31. I feel my body is a safe place. |  | TR |
| 24. When I bring awareness to my body, I feel a sense of calm. | 0.42 | SR |
| 25. I can use my breath to reduce tension. |  | SR |
| 24. When I bring awareness to my body, I feel a sense of calm. | 0.47 | SR |
| 26. When I am caught up in my thoughts, I can calm my mind by focusing on my body/breathing. |  | SR |
| 25. I can use my breath to reduce tension. | 0.62 | SR |
| 26. When I am caught up in my thoughts, I can calm my mind by focusing on my body/breathing. |  | SR |
| 27. I listen for information from my body about my emotional state. | 0.37 | BL |
| 28. When I am upset, I take time to explore how my body feels. |  | BL |
| 28. When I am upset, I take time to explore how my body feels. | 0.50 | BL |
| 30. I am at home in my body. |  | TR |
| 28. When I am upset, I take time to explore how my body feels. | 0.48 | BL |
| 31. I feel my body is a safe place. |  | TR |
| 30. I am at home in my body. | 0.73 | TR |
| 31. I feel my body is a safe place. |  | TR |
| 30. I am at home in my body. | 0.43 | TR |
| 32. I trust my body sensations. |  | TR |
| 31. I feel my body is a safe place. | 0.53 | TR |
| 32. I trust my body sensations. |  | TR |

4 Legend: RC (Residual Correlation), NOT (Noticing), ND (Not-Distracting), NW (Not- 5 Worrying), AR (Attention Regulation), EA (Emotional Awareness), SR (Self- 6 Regulation), BL (Body Listening), TR (Trusting)

